# Can motion capture improve task-based fMRI studies of motor function post-stroke? A systematic review

**DOI:** 10.1101/2024.03.24.24304778

**Authors:** Zakaria Belkacemi, Liesjet E.H. van Dokkum, Andon Tchechmedjiev, Matthieu Lepetit-Coiffe, Denis Mottet, Emmanuelle Le Bars

**Affiliations:** Siemens Healthineers France, Courbevoie, France; Euromov Digital Health in Motion, University of Montpellier, IMT Mines Alès, Montpellier, France; Institut d’Imagerie Fonctionnelle Humaine, Neuroradiology Department, Gui de Chauliac, Montpellier University Hospital, Montpellier, France

**Keywords:** fMRI, Motor function, Motion capture, Kinematics, Stroke, Multimodal, Neuroimaging, Review

## Abstract

**Background:** Variability in motor recovery after stroke represents a major challenge in its understanding and management. While functional MRI has traditionally been used to address post-stroke motor function in relation to clinical outcome, it lacks details about movement characteristics linked to observed brain activations. Combining fMRI with detailed information of motor function by using motion capture (mocap) might provide clinicians with additional information about mechanisms of motor impairment after stroke.

**Objectives:** We aimed to identify fMRI and mocap coupling approaches and to evaluate their potential contribution to the understanding of motor function post-stroke.

**Method:** A systematic literature review was performed according to PRISMA guidelines, on studies using fMRI and mocap in post-stroke individuals. We assessed the internal, external, statistical, and technological validity of each study. Data extraction included study design and analysis procedures used to couple brain activity with movement characteristics.

**Results:** Of the 404 studies found, 23 were included in the final review. The overall study quality was moderate (0.6/1). The majority of studies focused on the upper limb, using a wide variety of motor tasks. Half of the studies performed a statistical analysis between movement and brain activity by either using kinematics as variables during group or individual level regression or correlation. This permitted establishing a link between motor characteristics and brain activations. Mocap was also integrated without statistical confrontation, to compare results between fMRI and kinematics, or to incorporate real-time movement information to supply external devices, like motor feedback.

**Conclusion:** Our review suggests that the simultaneous use of fMRI and Mocap provides new insights compared with conventional fMRI analysis. It allows a better understanding of post-stroke motor function, although being still subject to practical and technological limitations. Further research is needed to optimize and standardize both data measurement and processing procedures.

## Introduction

Stroke is the second leading cause of death and disability worldwide, affecting more than ten million people worldwide each year (1). Among survivors, more than 60% show sequels like language, motor or cognitive disorders, which makes stroke a major public health problem (1). For example, a large study found that 51% of stroke survivors were unable to walk independently just after their stroke. After rehabilitation this amount decreased to 18% (2). In contrast, upper limb dexterity is less frequently recovered, with some dexterity retrieved in 38% of the cases and complete functional recovery in only 11.6% after six months of rehabilitation (3). Indeed, through rehabilitation, recovery of motor function is esteemed to be driven by brain plasticity, or the capacity of the brain to adapt itself after a lesion (4). However, the exact relation between brain plasticity and motor recovery is still subject to discussion (5,6). Brain plasticity can be studied via neuroimaging technologies (7) like electroencephalography (EEG), magnetoencephalography (MEG), functional Near Infrared Spectroscopy (fNIRS), or Positron Emission Tomography (PET). Amongst these, functional Magnetic Resonance Imaging (fMRI), that quantifies brain activity based on neurovascular coupling, has become the corner-stone in post-stroke research, with its high spatial resolution and a continuously improving temporal resolution (8). It has revealed global brain activity patterns that correlate with motor function (9), and has consequently improved our understanding of post-stroke motor recovery (10,11). Moreover, longitudinal fMRI studies highlighted the joint evolution of brain plasticity and motor recovery, like for instance the positive link between improved motor function and the return to normal brain activity patterns (10).

Nevertheless, the large variability in the amount of motor recovery after stroke leaves many questions open, like how to facilitate brain plasticity to optimize recovery for each individual patient (12). While the evolution of motor task-related brain activity has been clearly linked to recovery outcome as measured by clinical scales (13), only few studies investigated brain activity in relation to the characteristics of the performed motor task itself. In a neuroimaging review on upper-limb recovery after stroke, Buma et al. (10) highlight the need to control for task-related confounding factors during fMRI, especially in relation to the quality of task performance. They suggest controlling the execution of motor tasks to improve the understanding of the association between brain activity patterns and recovery. Indeed, without appropriate information on how the movement is performed within the fMRI, imaging data cannot distinguish whether changes in brain activity reflect neurological recovery or behavioral compensation (14). To improve understanding of brain plasticity, it has therefore been recommended to combine longitudinal task-related imaging with standardized analysis of the task performance. The most fine-grained manner to obtain such information is by means of a kinematic analysis, or, the study of motion (14). Kinematic analysis permits the characterization of the motor task in time and space, using a motion capture device. There are numerous kinematic parameters that quantify movement execution, and have been shown informative of healthy motor control as well as post-stroke (15). It has been shown that kinematics are better able to discriminate between different levels of post-stroke motor impairment than the Fugl-Meyer Assessment (FMA) (16), which has been the gold-standard to assess post-stroke motor impairment in rehabilitation research (17). Studies coupling functional neuroimaging with kinematics may thus provide further information on brain plasticity and the underlying motor control after stroke (18–22). Being a relatively novel field, this systematic review aims at analyzing the different approaches currently used and their related findings to identify the potential value of a combined task-fMRI and kinematic approach to study motor function after stroke.

## Materials and Methods

The systematic review meets the Preferred Reporting Items for Systematic Reviews and Meta-Analysis (PRISMA) requirements (23).

### Inclusion Criteria

Inclusion criteria were English written, full-text studies using motor task fMRI of the upper or lower extremity after stroke, with kinematic assessment of the motor task by motion capture, regardless the type of motion capture device. All published studies and preprints meeting the inclusion criteria until August 2022 were included. Reviews and conference abstracts were excluded.

### Search Strategy

The literature search was performed by two authors (ZB and LvD) and supported by a third author in case of discussion (ELB). The following search terms were added to Medline, Embase, Web of Science, and IEEE Explore: ((fMRI) OR (functional magnetic resonance imaging) OR (functional neuroimaging)) AND (Stroke) AND ((motor control) OR (movement)) AND ((motion tracking) OR (motion capture) OR (kinematics) OR (movement smoothness) OR (motion analysis)). We did not use automatic tools to also include papers in which the kinematics coupled with fMRI approach appeared as a secondary objective.

### Assessment of Methodological Quality of studies

Methodological quality was assessed with an adapted version of the clinical methodological rounds (24) following Buma et al. (10), who systematically reviewed serial imaging studies to identify trends in the association between brain activity and functional upper limb recovery after stroke. To comply with our objective of analyzing the value of a combined fMRI/kinematics approach, items evaluating the internal, statistical, and external validity were modified accordingly. A fourth scale was also added to evaluate the technological validity of each study. The criteria of internal validity were broadened to include both lower and upper-limb studies, whether cross-sectional or longitudinal, and limited to imaging by means of fMRI. We also added a criterion to the statistical validity, covering the integration of kinematics in the fMRI statistical analysis. The resulting Methodological Quality Assessment with a short description of each item is shown in table 1. A detailed description of each item can be found below.

**Table 1:** Methodological Quality Assessment.

| Item | Description |
| --- | --- |
| **Internal validity** |  |
| 1: Measurements of motor function | Positive if measurement of motor function is effectuated with clinically relevant and validated tests. |
| 2: Clear presentation of fMRI parameters | Positive if MRI parameters are clearly described in the methods section. |
| 3: Description of additional medical and paramedical interventions | Positive if information on medical and paramedical treatment is reported. |
| 4: Mirror movement assessment | Positive if mirror movements during the fMRI session are assessed with e.g., EMG, kinematics, or visual inspection. |
| 5: Control of motor task performance | Positive if movement amplitude, frequency or range of motion are either standardized or measured during task execution. |
| **Statistical validity** |  |
| 6: Multiple comparisons correction | Positive if a correction for multiple comparisons has been applied to P-values for brain activity. |
| 7: Validity of applied statistics within and between subjects | Positive if applied statistical analyses within and between subject analyses are appropriate to the population and the study design |
| 8: Combined fMRI and kinematic analysis | Positive if statistical tests are performed between fMRI and kinematic data. |
| **External validity** |  |
| 9: Specification of relevant patient characteristics | Positive if age, type of stroke, location, and number of strokes are specified. |
| **Technological validity** |  |
| 10: MRI strength | Positive if the strength of the magnetic field in Tesla is $\geq 3$. |
| 11: fMRI spatial resolution | Higher scores correspond with higher spatial resolution of the fMRI sequence. |
| 12: fMRI temporal resolution | Higher scores correspond with a higher temporal resolution (TR: repetition time) of the fMRI sequence. |
| 13: Constraining character of the motion capture device. | The lower the impact of the motion capture device on the ecological nature of the movement, the higher the score. |

Detailed description of the items of the methodological quality assessment:

1. Motor function (0-1 point): Measurement of motor function had to be assessed with validated measures like the Fugl-Meyer-Assessment of the Upper Extremity (FMA-UE) (17), Box and Block Test (BBT) (25), Nine-hole Peg Test (NHPT) (26), Action Arm Reach Test (ARAT) (27) or Wolf Motor Function Test (WMFT) (28) for the upper-limb, or with the 50-feet walking test (29), 10-meter walking test, 6-minutes walking test, Motricity Index of the Lower-Limb or the Fugl-Meyer Assessment of Lower Extremity test (29,30) for the lower-limb.
2. Presentation of fMRI parameters (0-1 point): Positive if fMRI parameters are clearly described: pre- and post-processing procedures, statistical analysis including cluster size and location, software, and brain atlas used.
3. Description of additional medical or paramedical interventions (0-1 point): Positive if the study reports the verification of additional medical or paramedical interventions which might have an impact on fMRI results (e.g., treatment with botulinum toxin).
4. Mirror Movement assessment (0-1 point): Positive if the study controls for mirror movements with the contralateral limb, assessed with either EMG, kinematics, or visually during unilateral motor tasks. Mirror movements of the contralateral limb during paretic limb activity biases the corresponding activity patterns and should be taken into account in the analysis (31).
5. Motor task monitoring (0-1 point): Positive when movement pace and amplitude are either fixed or monitored, because they impact the intensity of the BOLD signal (32,33).
6. Correction for multiple comparisons (0-1 point): Positive if P-values for activated brain areas are corrected for multiple comparisons, for example, by applying a Bonferroni correction or a Family Wise Error (FWE) correction.
7. Applied statistics (0-1 point): Positive if the applied statistics for within and between subject analyses are in accordance with the number of participants and the question addressed by the test.
8. Combined kinematics and fMRI analysis (0-1 point): Positive if kinematic and fMRI results are confronted using a statistical approach.
9. Specification of patient information (0-1 point): Positive if age, type, location, and number of strokes are specified.
10. MRI strength (0-1 point): Higher magnetic field strengths improve the measurement of the BOLD response (34,35). The MRI strength is considered positive if the magnetic field is superior or equal to 3 Tesla.
11. fMRI spatial resolution (0 - 0.33 - 0.67 - 1 point): The precision of fMRI results increases with increasing spatial resolution (36). The spatial resolution is defined by the transverse resolution plane (x,y) in mm, and the slice thickness (z) in mm. The following gradation has been applied:

- 0 point if both x and y > 4
- 0.33 point if both x and y ∈ [3:4]
- 0.67 point if both x and y ∈ [1:2] and z > 2
- 1 point if both x and y ∈ [1:2] and z ≤ 2
12. fMRI temporal resolution (0 - 0.33 - 0.67 - 1 point): Higher temporal resolution reduces physiological noise and the under sampling effect, increasing the SNR efficiency [29] and providing a better sensitivity (37). The following gradation has been applied:

- 0 point if TR > 3 seconds
- 0.33 point if TR ∈ [2:3] seconds
- 0.67 point if TR ∈ [1:2 [seconds
- 1 point if TR ∈ [0.5:1] second
13. Motion Tracking device (0.3 - 0.6 - 1 point): To evaluate and quantify kinematics, various devices exist with more or less impact on the ecological character of the movement, or, whether the movement can be performed as natural as possible. For example, haptic gloves alter the sensory feedback of a movement and thus the way the movement is controlled (38). Such devices are considered <u>highly constraining</u> and little representative of ecological motion. Subsequently, studies using non wireless devices or devices that are strapped to the participant are considered <u>constraining</u>. In contrast, wireless small markers for optical motion tracking interfere only slightly with the natural/ecological movement and are considered <u>slightly</u> <u>constraining.</u>

- 0 point if highly constraining (robot, haptic glove, goniometer)
- 0.5 point if constraining (non-wireless, large markers, strapped devices)
- 1 point if slightly constraining (wireless, small markers ≤ 15mm diameter)

### Data collection

The following additional information was extracted from each study to identify study characteristics:

- The type of the study, e.g., a longitudinal, a cross-sectional, or a pilot-study.
- The aim of the study, e.g., evaluation of a therapeutic approach, understanding brain function, or a technological proof of concept.
- The calculated kinematics, or the amount and nature of the kinematic parameters assessed.
- The recording conditions: e.g., the time of kinematic acquisition (during the fMRI acquisition or not).
- The type of statistical analysis that was performed between brain imaging and kinematics.
- The results of the brain imaging with kinematic analysis.

## Results

### Literature search

The search terms yielded 404 papers, including 207 from Medline, 56 from Embase, 141 in Web of Science and 2 in IEEE Explore (details are listed in Appendix A). Sixty-six papers were retrieved based on title and abstract screening, dismissing papers that did not respect inclusion criteria (figure 1). The full in-depth evaluation of these papers led to the final inclusion of 23 studies (18,19,19–22,39–56), excluding those that were about fMRI alone, kinematics alone or that did not involve people post-stroke. No additional papers were found by citation.

**Figure 1:**
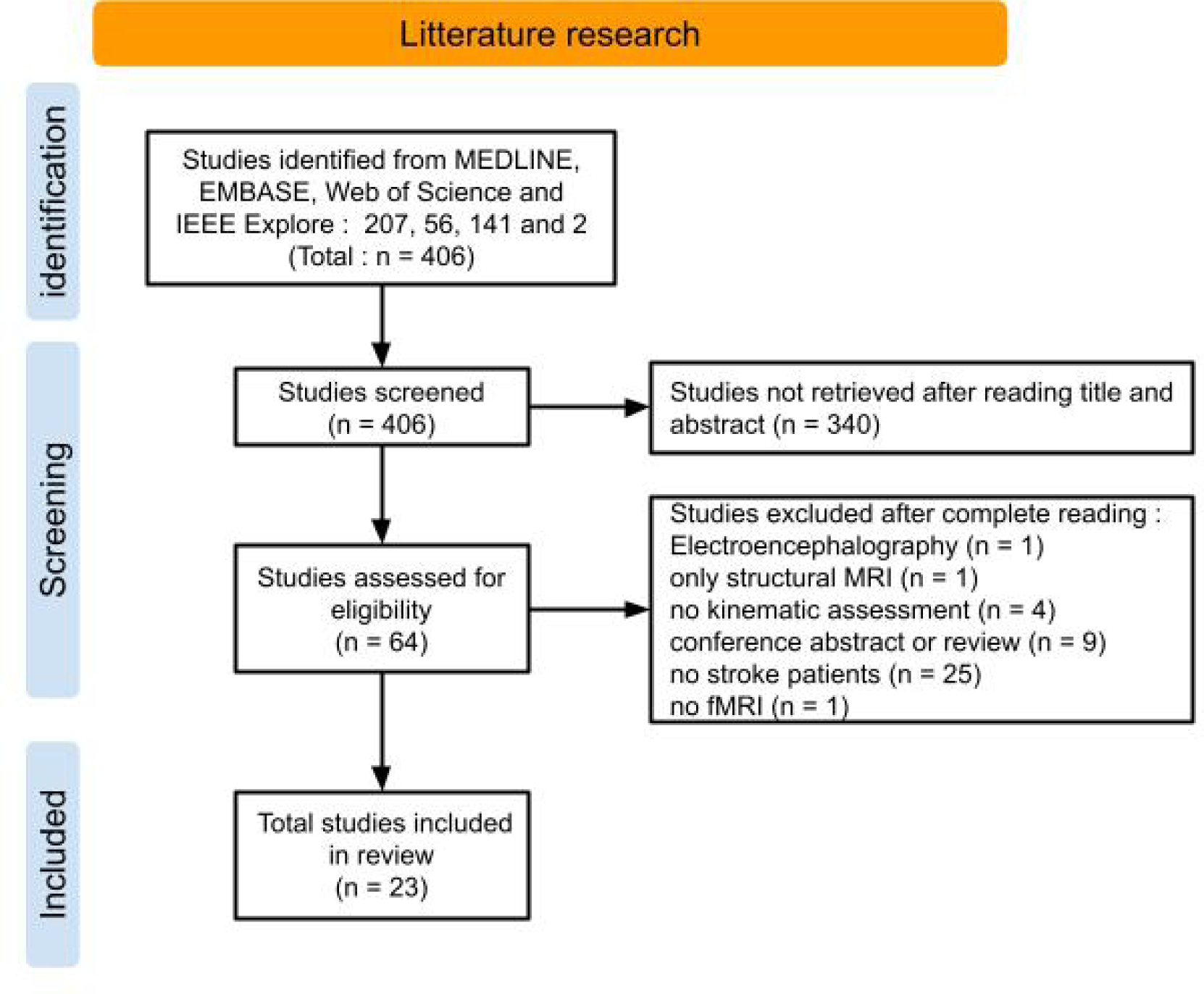
Flow-chart of study selection.

### Methodological Quality of studies

The methodological quality varied largely over studies ranging from 0.28 to 0.69. Still, with a mean score of 0.66 the overall quality of the included studies was found to be sufficient. Especially technological and statistical validity present themselves as the weakest areas of validity, with respectively a mean quality of 0.48 and 0.56 (figure 2). Because of the limited sample-size and the heterogeneity of studies objectives, we did not set a quality-based exclusion threshold. An overview of the methodological quality assessment results can be found in table 2.

**Figure 2:**
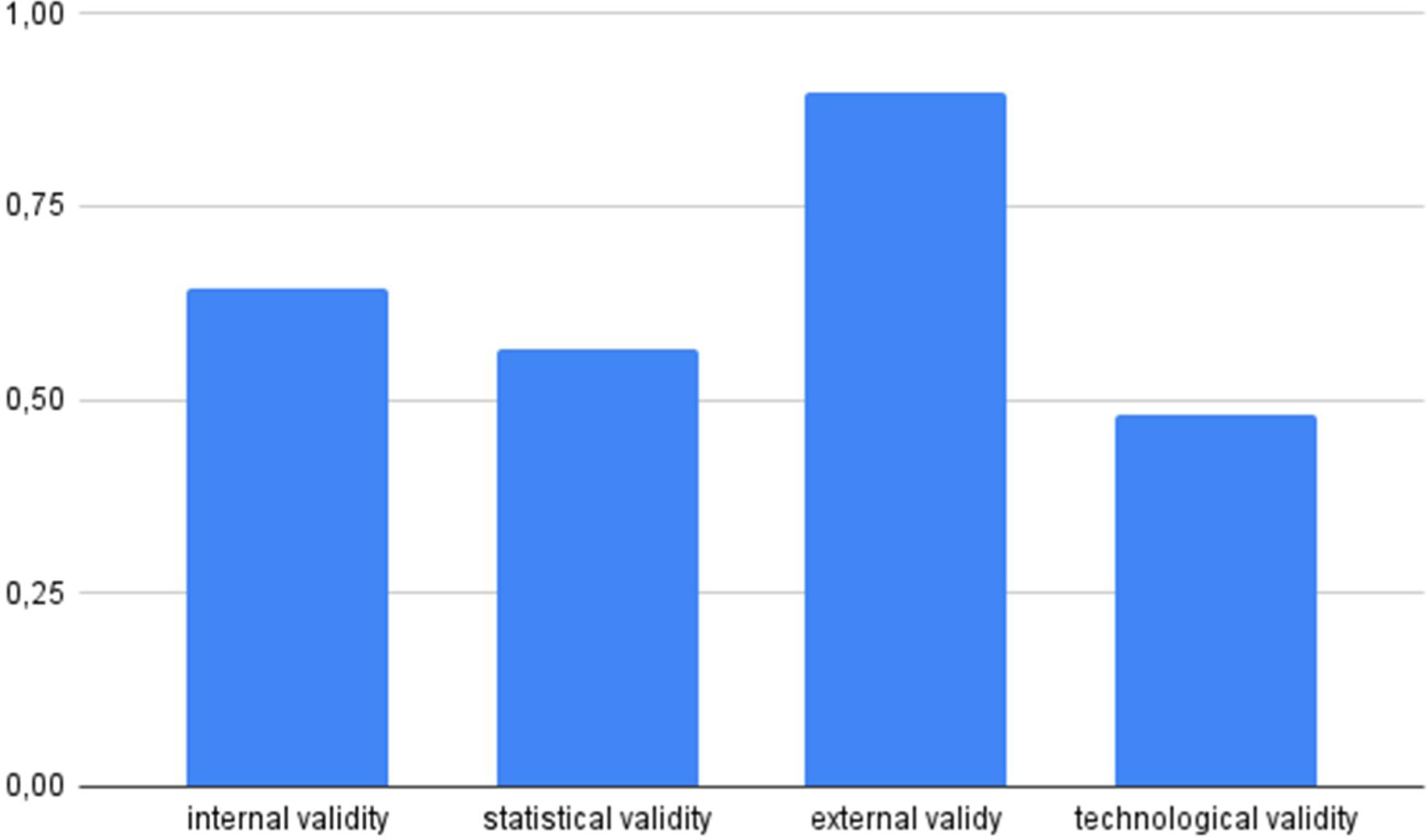
Mean score for each type of validity reported between 0 and 1. Validities refer to categories of items described in the Methodological quality assessment (Table 1).

**Table 2:** Methodological Quality Assessment Scores.

| Methodological Quality |  |  |  |  |  |  |  |  |  |  |  |  |  |  |  |  |
| --- | --- | --- | --- | --- | --- | --- | --- | --- | --- | --- | --- | --- | --- | --- | --- | --- |
| Validity | Int |  |  | Stat |  |  | Ext |  |  |  | Tech |  |  |  |  |  |
| Item | 1 | 2 | 3 | 4 | 5 | 6 | 7 | 8 | 9 |  | 10 | 11 | 12 | 13 |  |  |
| Full Item name →<br>Reference nr + first author ↴ | Motor function measurement | fMRI processing description | Additional intervention description | Mirror Movement assessment | Motor task monitoring | Multiple comparison correction | Statistics | Use of Kinematics in fMRI analysis | Specification of patient information | Methodological Quality | MRI strength | fMRI spatial resolution | fMRI temporal resolution | Motion Tracking device constraint | Technological Quality | Mean of Methodological Technological Quality |
| (39) Ameli 2009 | 1 | 1 | 0 | 0 | 0 | 1 | 1 | 1 | 1 | 0.67 | 1 | 0.33 | 0.67 | 0.5 | 0.63 | 0.65 |
| (40) Bani-Ahmed 2020 | 0 | 1 | 1 | 1 | 1 | 0 | 1 | 1 | 1 | 0.78 | 1 | 0 | 0 | 1 | 0.5 | 0.64 |
| (18) Brihmat 2020 | 0 | 1 | 0 | 0 | 1 | 1 | 1 | 1 | 0 | 0.56 | 1 | 0.33 | 0.33 | 0 | 0.42 | 0.49 |
| (22) Buma 2016 | 1 | 1 | 1 | 1 | 1 | 1 | 0 | 0 | 1 | 0.67 | 1 | 0.33 | 1 | 0.5 | 0.71 | 0.69 |
| (42) Carey 2007 | 1 | 1 | 0 | 1 | 1 | 0 | 1 | 0 | 1 | 0.67 | 1 | 0.33 | 0.33 | 0 | 0.42 | 0.55 |
| (41) Carey 2004 | 1 | 1 | 0 | 1 | 1 | 0 | 0 | 0 | 1 | 0.44 | 1 | 0.33 | 0.33 | 0 | 0.42 | 0.43 |
| (19) Casellato 2010 | 1 | 1 | 0 | 1 | 1 | 1 | 1 | 1 | 1 | 0.89 | 0 | 0.67 | 0.33 | 1 | 0.5 | 0.7 |
| (43) Ciceron 2022 | 1 | 1 | 0 | 1 | 1 | 1 | 0 | 1 | 1 | 0.78 | 0 | 0.33 | 0.33 | 1 | 0.42 | 0.6 |
| (20) Del Din 2014 | 1 | 1 | 0 | 0 | 1 | 0 | 0 | 0 | 1 | 0.44 | 0 | 0.33 | 0.33 | 1 | 0.42 | 0.43 |
| (44) Deng 2012 | 1 | 1 | 0 | 1 | 1 | 1 | 1 | 0 | 1 | 0.78 | 1 | 0.33 | 0.33 | 0 | 0.42 | 0.6 |
| (45) Dobkin 2004 | 1 | 1 | 0 | 1 | 1 | 0 | 1 | 0 | 0 | 0.56 | 0 | 0.33 | 0.33 | 0.5 | 0.29 | 0.43 |
| (46) Gandolla 2021 | 1 | 1 | 0 | 0 | 1 | 0 | 1 | 0 | 1 | 0.56 | 0 | 0.67 | 0.33 | 1 | 0.5 | 0.53 |
| (47) Hensel 2021 | 1 | 1 | 0 | 0 | 1 | 1 | 1 | 0 | 1 | 0.67 | 1 | 1 | 1 | 0.5 | 0.88 | 0.78 |
| (48) Hensel 2023 | 1 | 1 | 0 | 0 | 1 | 1 | 1 | 0 | 1 | 0.67 | 1 | 1 | 1 | 0.5 | 0.88 | 0.78 |
| (49) Nowak 2008 | 1 | 1 | 0 | 0 | 0 | 1 | 0 | 1 | 1 | 0.56 | 1 | 0.33 | 0.67 | 0 | 0.5 | 0.53 |
| (50) Promjunyakul 2015 | 1 | 1 | 0 | 0 | 1 | 0 | 1 | 1 | 1 | 0.67 | 1 | 0.33 | 0.33 | 0 | 0.42 | 0.55 |
| (51) Saleh 2011 | 1 | 1 | 0 | 1 | 1 | 0 | 0 | 1 | 1 | 0.67 | 1 | 0.33 | 0.33 | 0 | 0.42 | 0.55 |
| (21) Saleh 2014 | 1 | 1 | 1 | 1 | 1 | 1 | 1 | 0 | 1 | 0.89 | 1 | 0.33 | 0.33 | 0 | 0.42 | 0.66 |
| (52) Schaechter 2008 | 1 | 1 | 0 | 1 | 1 | 0 | 1 | 0 | 1 | 0.67 | 1 | 0.33 | 0.67 | 0.5 | 0.63 | 0.65 |
| (53) Sergi 2011 | 0 | 1 | 0 | 0 | 0 | 1 | 1 | 0 | 1 | 0.44 | 1 | 0.33 | 0.33 | 0.5 | 0.54 | 0.49 |
| (54) Tunik 2013 | 0 | 1 | 0 | 0 | 0 | 0 | 0 | 0 | 1 | 0.22 | 1 | 0.33 | 0.33 | 0.5 | 0.54 | 0.38 |
| (55) Turolla 2013 | 1 | 1 | 1 | 0 | 1 | 0 | 0 | 0 | 0 | 0.44 | 0 | 0 | 0.33 | 0 | 0.08 | 0.26 |
| (56) van Dokkum 2018 | 1 | 1 | 0 | 0 | 1 | 1 | 1 | 1 | 1 | 0.78 | 0 | 0.33 | 0 | 0.5 | 0.21 | 0.5 |
| Mean Score | 0.8 | 1 | 0.2 | 0.5 | 0.8 | 0.5 | 0.7 | 0.4 | 0.9 | 0.63 | 0.7 | 0.4 | 0.4 | 0.4 | 0.5 | 0.56 |

#### Internal validity

Nineteen studies measured upper and/or lower-limb motor function with clinically relevant and validated tests (19–22,39,41–52,55,56). Considering the upper-limb, the ARAT was the most used measure with five occurrences. Four studies used the BBT, two studies used the WMFT, and only one study used the NHPT as a measure of initial upper-limb motricity. Among the seven lower-limb studies, the walking speed was used five times to assess motricity. In addition, Casellato et al. (11) used the Motricity Index for the Lower Limb, and Huiquiong used the ten-meter walk test. Other gait parameters were the stride length or symmetry ratio between the two legs during walking. Only four studies (21,22,40,55) reported medical or drug conditions in patients that could have interfered with the functional MRI results. In all cases they controlled either for post-stroke spasticity treatment with botulinum toxin injection or for hypertension treatment. Eleven studies took into account potential mirror movements (19,21,22,40–45,51,52), by using electromyogram (EMG) (22), visual inspection (40,41,43,45), or motion capture (19,21,42,44,51,52). The movement frequency was paced in nine studies with either an auditory or visual signal (18,22,40–42,44–46,49,52,54) (18,22,40– 42,45,46,49,52). Four studies constrained the movement amplitude with an orthosis, a cast, or a brace (42,45,53,55). One study was paced and constrained in amplitude(45). Meanwhile nine studies used a free movement with no pacing and without fixing the body member of interest (19–21,39,43,47,48,50,56), but seven of them controlled the amplitude thanks to a motion tracking device (19,21,50,56) or by visual control (43,47,48). The majority of studies evaluated brain activity using a block design fMRI protocol, signified by alternating periods of continuous movement with periods of rest (18–20,22,39–45,47–50,52,55,56). Only five studies used an event-related design, defined by the repeated execution of one distinct task at certain defined times (21,46,51,53,54).

#### Technological validity

For the kinematic assessment, five studies used optical motion capture and were classified as “slightly-constraining” (19,20,40,43,46). Seven studies were classified as “constraining”, among which one used electromagnetic motion tracking system (22), five used ultrasonic (39,47–49,56), and two used accelerometers (45,52). Finally, nine studies were classified as “very constraining”, among which two used data-gloves (21,54), two used a rehabilitation robot (51,53), four used a data-goniometer (18,41,42,44) and one used a custom-made leg-press recorder (50).

### Data collection

In the following section, we describe the extracted additional information related to: the study design, the functional task, the kinematic parameters assessed, and the joint analysis performed between fMRI and kinematic data.

#### Study design

Among the twenty-three studies, eleven were longitudinal studies (19,22,40–42,45,46,49,51,55,56), mostly evaluating motor recovery in a pre/post rehabilitation design. Twelve studies were cross-sectional (18,20,21,39,43,44,47,48,50,52–54). And five studies were pilot, case or feasibility studies, which were either longitudinal or cross-sectional in character (19,20,41,43,55). The main objective of fifteen studies was the analysis of brain activity patterns, while four studies focused on the evaluation of a rehabilitation program. In addition, there were two feasibility studies evaluating the integration of an MRI-compatible kinematic system, and one study was about the prediction of rehabilitation efficiency.

#### Motor task configuration

The majority of studies were interested in motor function of the upper-limb (17/23) with a variety of functional tasks performed during fMRI, including finger flexion (21,22,42,51,52,55), wrist flexion (18), elbow flexion (56), finger tapping (19,39,47,48), hand tapping (39), finger opposition (43), and handgrip (40,49). Five studies included a reach to grasp task that was performed outside of the MRI (22,40,43,48,49). The seven studies interested in the lower limb used either a pedaling (50) or ankle flexion task (19,20,41,44–46).

#### Kinematic parameters

To get an overview of the type of kinematic parameters used, we regrouped all kinematic parameters that were used within the seven domains described by Schwarz et al. 2019 (15), notably ‘efficiency’, ‘speed’, ‘smoothness’, ‘temporal posture’, ‘planning’, ‘accuracy’, and ‘spatial posture’ (figure 3). More than fifty percent of the kinematics covered the efficiency and speed domain. The efficiency domain was mainly represented by kinematic parameters that described the execution time and the movement amplitude. The speed domain was represented by both movement velocity and frequency measures.

**Figure 3:**
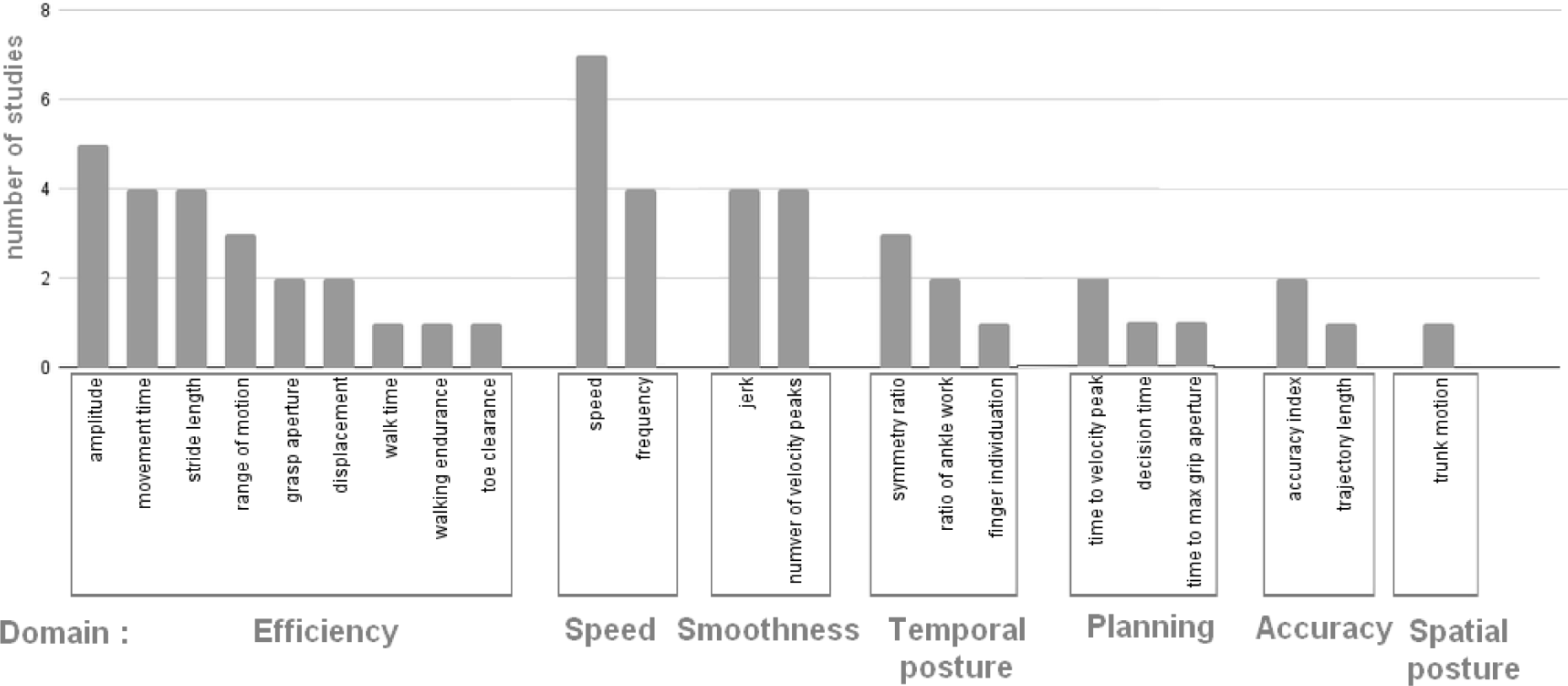
Number of studies per kinematic parameter, regrouped within seven domains as defined by Schwarz et al. 2019 (15).

#### Brain and movement analysis

Thirteen studies recorded fMRI and motion capture simultaneously (18,19,21,41,42,44,46,50–54,56). The other ten studies used a non-simultaneous tracking in which motion capture of a comparable task was performed outside of the MRI. An overview of all collected data can be found in table 3.

**Table 3.**

| Reference | Study design |  |  |  | Motor task |  |  | Analysis |  |  |
| --- | --- | --- | --- | --- | --- | --- | --- | --- | --- | --- |
|  | study type | study aim | number of participants |  | within MRI | outside MRI | limb studied | Mocap during fMRI? | Kinematic parameters | Use of kinematics in fMRI |
| (39)<br>Ameli<br>2009 | cross-sectional | brain study | 21 | 0 | index finger tapping | index finger & hand tapping | upper | no | tapping frequency | covariable in group analysis and response identification to repetitive TMS |
| (40)<br>Bani-Ahmed<br>2020 | longitudinal | brain study | 11 | 12 | handgrip | reaching | upper | no | trunk movement | covariable in group analysis |
| (18)<br>Brihmat<br>2020 | cross-sectional | brain study | 15 | 0 | passive wrist extension |  | upper | yes | amplitude | covariable in group analysis and regressor in individual analysis |
| (22)<br>Buma<br>2016 | longitudinal | brain study | 15 | 0 | finger flexion | reaching | upper | no | grasp aperture, normalized jerk | group correlation with BOLD signal in Regions of Interest |
| (42)<br>Carey<br>2007 | longitudinal | rehabilitation evaluation | 20 | 0 | paretic index finger flexion |  | upper | yes | tracking accuracy, range of motion | visual feedback during task execution |
| (41)<br>Carey<br>2004 | longitudinal case study | rehabilitation evaluation | 1 | 0 | Unilateral ankle flexion |  | lower | yes | accuracy index, walking time, ankle range of motion, peak dorsiflexion | comparison of results |
| (19)<br>Casellato<br>2010 | longitudinal pilot study | feasibility and brain study | 1 | 1 | ankle flexion, finger tapping |  | lower & upper | yes | angular amplitude, frequency, between feet correlation, displacement | regressor in individual analysis |
| (43)<br>Ciceron<br>2022 | cross-sectional case study | brain study | 1 | 10 | finger opposition | reaching | upper | no | movement time, peak velocity, time to peak velocity, maximal grip aperture, time to maximal grip aperture | to distinguish motor recovery from motor compensation |
|  | study type | study aim | nr pp |  | within MRI | outside MRI | limb studied | Mocap during fMRI? | Kinematic parameters | Use of kinematics in fMRI |
| (20)<br>Del Din<br>2014 | cross-sectional<br>case study | brain study | 1 | 1 | ankle flexion | gait | lower | no | cadence, stride length, peak power, positive/negative work | correlation at the individual level with BOLD signal |
| (44)<br>Deng<br>2012 | cross-sectional | rehabilitation evaluation | 15 | 0 | ankle flexion | gait | lower | yes | dorsiflexion angle, toe clearance, symmetry ratio, stride length | verification of mirror movements |
| (45)<br>Dobkin<br>2004 | longitudinal | brain study | 1 | 12 | ankle flexion | gait | lower | no | walking speed | evaluation of motor evolution between training sessions |
| (46)<br>Gandolla<br>2021 | longitudinal | brain study | 8 | 16 | right active & passive ankle flexion | gait | lower | yes | gait velocity, endurance velocity, paretic step length | monitoring Functional Electrical Stimulation |
| (47)<br>Hensel<br>2021 | cross-sectional | brain study | 14 | 13 | finger tapping |  | upper | no | peak velocity | guiding Transcranial Magnetic Stimulation |
| (48)<br>Hensel<br>2023 | cross-sectional | brain study | 18 | 18 | finger tapping | finger tapping, pointing, reaching | upper | no | efficiency, accuracy, smoothness, speed | correlation with connectivity |
| (49)<br>Nowak<br>2008 | longitudinal | brain study | 15 | 0 | handgrip | tapping, reaching | upper | no | time of peak velocity, peak velocity | covariable in group analysis |
| (50)<br>Promjunyakul<br>2015 | cross-sectional | feasibility study | 14 | 12 | pedaling |  | lower | yes | step length, walking velocity, symmetry, work ratio paretic/non-paretic side | correlation with BOLD signal |
|  | study type | study aim | nr pp |  | within MRI | outside MRI | limb studied | Mocap during fMRI? | Kinematic parameters | Use of kinematics in fMRI |
| (51)<br>Saleh<br>2011 | longitudinal | brain study | 4 | 0 | finger flexion |  | upper | yes | angular velocity, smoothness, finger individuation, range of motion | correlation with BOLD signal |
| (21)<br>Saleh<br>2014 | cross-sectional | brain study | 15 | 0 | finger flexion |  | upper | yes | movement time, mean peak angular velocity | visual feedback during task execution |
| (52)<br>Schaechter<br>2008 | cross-sectional | brain study | 10 | 10 | synergistic & non-synergistic digits flexion |  | upper | yes | amplitude, frequency, speed, acceleration, jerk, mirroring | comparison of results |
| (53)<br>Sergi<br>2011 | cross-sectional | rehabilitation efficacy prediction | 2 | 2 | reaching |  | upper | yes | velocity, movement duration, displacement | analysis of kinematics alone |
| (54)<br>Tunik<br>2013 | cross-sectional | brain study | 3 | 12 | sequential finger movement |  | upper | yes | movement duration, mean displacement, decision time | visual feedback during the task, and use of kinematic data to confirm that subjects complied with the task |
| (55)<br>Turolla<br>2013 | longitudinal pilot study | rehabilitation evaluation | 1 | 0 | index flexion |  | upper | no | movement time, normalized jerk | comparison of results, evolution evaluation between sessions |
| (56)<br>van Dokkum<br>2018 | longitudinal | brain study | 19 | 13 | elbow flexion |  | upper | yes | amplitude, frequency, normalized trajectory length, number of velocity peaks | covariate in group analysis |

The timing of kinematic recording, during fMRI or not, was unrelated to the use of kinematics. We identified five different ways to use motion capture in a fMRI context. First, to facilitate the interpretation of the BOLD signal without any statistical analysis between kinematic and fMRI parameters (41,43,45,52,54,55). Secondly, to optimize the fMRI contrast paradigm by using kinematics to define the action and rest blocks at individual-scale (19). Third, to guide related therapeutic interventions like transcranial magnetic stimulation (47), Functional Electrical Stimulation (FES) (46). Fourth, to provide the participant with visual feedback (21,42,54). Fifth, to proceed to a statistical analysis, identifying brain regions that correlate with certain movement characteristics. The latter was achieved by integrating kinematic either as covariates at a group level fMRI analysis (18,39,40,49), or as regressors at an individual level fMRI analysis (18,19), or in a correlation with brain regions’ BOLD signals at the individual level (20,51), or at the group level (22,48). Note that only half of the studies that registered kinematics and fMRI simultaneously, performed a statistical analysis between both techniques. In contrast, all the studies that performed a coupled analysis at the level of the individual participant acquired kinematics during fMRI (figure 4).

**Figure 4:**
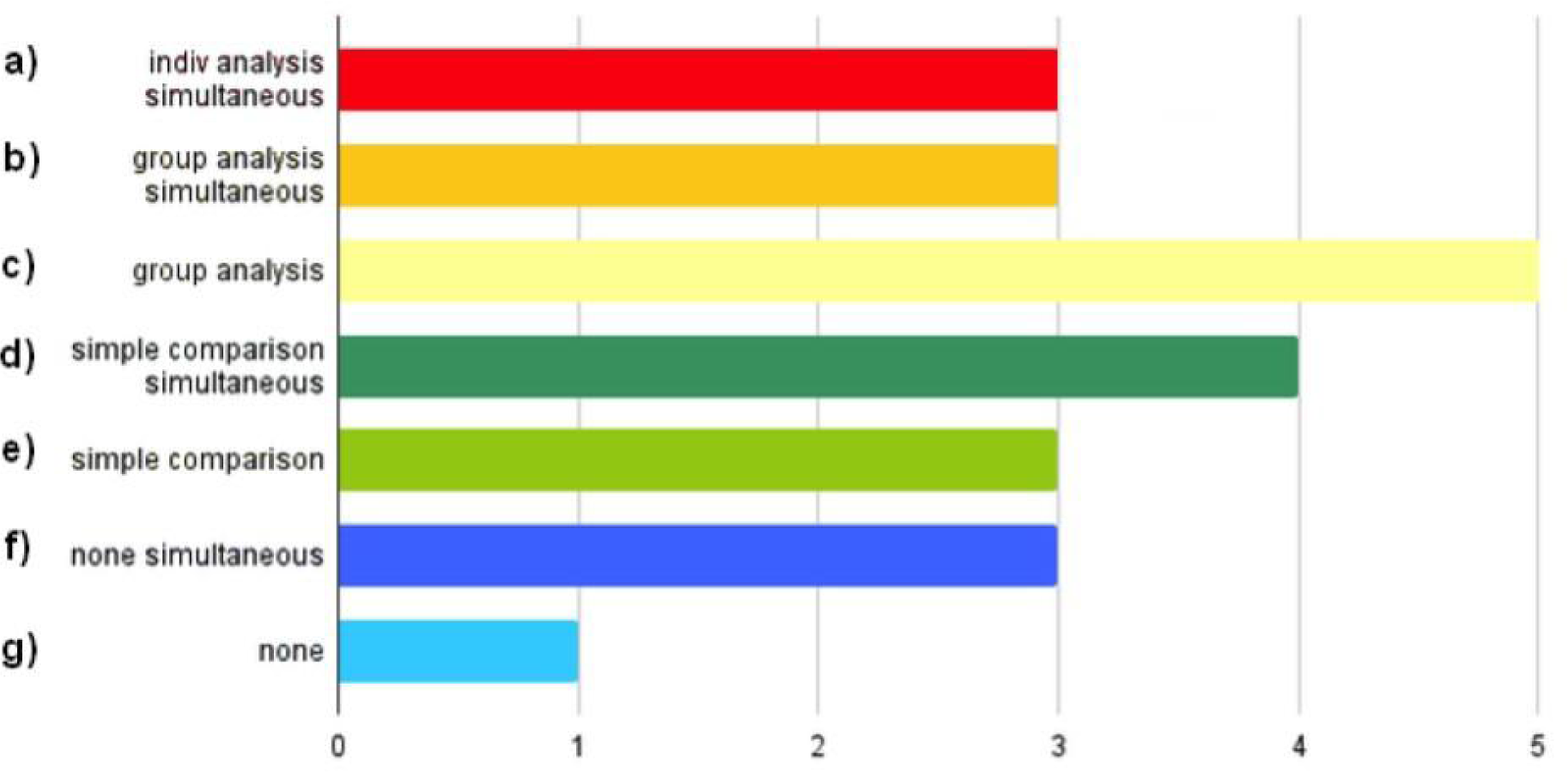
Use of Kinematics in the fMRI analysis. This figure indicates for the twenty-three studies, how mocap (motion capture) is used: (a) fMRI + kinematic analysis at the individual level, with a simultaneous mocap, (b) fMRI and kinematic analysis at the group level with a simultaneous or, (c) a non-simultaneous mocap, (d) a comparison of results between separately treated fMRI data and kinematics based on a simultaneous, or (e) a non-simultaneous mocap, (f) no statistical comparison between fMRI data and kinematics, although both have been recorded in a simultaneous, or (g) non simultaneous manner. A simultaneous capture is defined as the recording of the movement performed during the fMRI exam by a mocap device. Note that one study can score positive on multiple items.

By integrating kinematics at group level, Bani-Ahmed et al. demonstrated that the activity of the primary motor cortex (M1) during a hand-grip task varied with the amount of trunk displacement during a reaching task chronically post-stroke (40). Buma et al. demonstrated the additional recruitment of secondary sensorimotor areas as a function of finger flexion/extension smoothness (22). Ameli et al. found that baseline ipsilesional M1 activity correlated with the functional improvement in finger tapping frequency following repetitive TMS (39), whereas Nowak did not observe any correlation between rTMS-modified activity of the contralesional M1 during hand grip movements of the affected hand and its amount of functional improvement (49). Finally, Promjunyakul et al. were unable to identify any relationship between the lower-limb pedaling rate and the amount of brain activity (50).

Integrating kinematics at the individual level seems not to facilitate the identification of specific brain regions or networks, but rather improves the statistical significance and intensity of the task-related brain activity. For instance, Casellato et al. demonstrated a greater difference in brain activity levels between rest and movement periods in the fMRI block-design when using kinematics to identify when the participant was in motion and when at rest (19). Brihmat et al. showed that the amount of cerebellar activity decreased when the amplitude of the passive hand motion was used as a time series for individual level regression with brain BOLD signal (18). In contrast, studies using a longitudinal design that integrate kinematics at the individual level of the fMRI analyses, allow identifying changes in neuromotor coupling strength over rehabilitation. For instance, Saleh et al showed in two out of four participants increased correlation strength between ipsilesional sensorimotor activity and the angular velocity of finger flexion (51). Finally, in a case study design, Del Din et al demonstrated that the participant’s improvement in walking correlated with a greater and improved activation of the affected hemisphere (20).

## Discussion

In this review we looked for papers that combined brain imaging and kinematics to better understand motor function after stroke. For clarity, references concerning the reviewed papers are identified by a star (*) throughout the discussion. We were particularly interested in the novel information that could be gained by the specific combination of a joint fMRI and kinematic analysis. Twenty-three studies met our inclusion criteria, highlighting the novelty of the field on the one hand, and the technological complexity of both the integration of kinematic recording and analysis within fMRI, as emphasized by the presence of four technological feasibility studies *(19,20,55,43). Still, the global methodological quality was generally sufficient, although some studies lacked statistical power with poor internal validity, particularly when it came to taking into account potential interference with adjuvant medical interventions and the monitoring of mirror movements. Among each type of validity, technological validity is the weakest (with a mean of 0.48/1). This suggests a potential evolution on this point, since modern studies feature improved MRI power and spatial resolution. The statistical validity is the second weakest type of validity (0.56). This can be partly attributed to a large number of studies that did not integrate kinematics into the fMRI statistical analysis. Moreover, all works studied less than ten patients with stroke, which can compromise the correction for multiple comparisons. In the following we will first address the strengths and flaws of capturing movement in the fMRI, as well as the kinematic approaches used, before we will progress towards the coupled fMRI and kinematic analysis and its current and future challenges.

### fMRI and kinematics for both upper and lower-limb research?

Of the twenty-three studies, only seven studies addressed motor function of the lower limb. A potential explanation might be that the principal lower limb function is walking. As described in the introduction, most people after stroke recover independent walking with or without support. Locomotion is a specific motor function that is controlled at both the spinal and central level. Shortly stated, the spinal circuit generates the repetitive basic locomotion pattern, whereas the central descending pathways trigger, stop and steer locomotion (57) Making use of the often intact spinal circuits after stroke to stimulate central brain activity, locomotor rehabilitation practice has been well established by means of early treadmill training with body weight support that is progressively decreased with the improving gait pattern (58,59), with additional innovative interventions to improve walking speed and reduce spasticity, like brain computer interfaces (60) or non-invasive brain stimulation (61). Still, many people post-stroke, do not recover gait at a level sufficiently to participate in daily living activities like they used to, making lower-limb recovery still a subject of interest. And although walking cannot be simulated in an fMRI, elements of gait can. That is, ankle dorsiflexion, it represents an important kinematic aspect of the swing and initial stance phase of the gait cycle, and is identified as a practical substitute to address walking with fMRI (45). Correspondingly, most studies included, used an ankle dorsiflexion task to investigate gait.

In contrast to lower-limb recovery, upper-limb recovery is more complex as its main function is less specific, and more importantly, its functioning is fully organized at the central level. About 80% of stroke survivors experience upper limb impairments, and these movements have been well-researched in rehabilitation (62). Presumably due to the relative simplicity of recording these movements with limited risk of injury (unlike the falling-risk during gait) and the importance of such movements for daily activities. However, except for constraint induced therapy, no intervention has been proven more successful than another (63). Several explanations have been provided from inadequate targeting of motor control deficits to individual differences between patients, to functional task specificity and relevance (64). To yield the upper-limb rehabilitation field forward and improve rehabilitation gains, experts emphasize the need to quantify motor function in a standardized manner through kinematics and to increase the understanding of kinematics significance by MRI research (14). All these elements may underlie the larger interest of fMRI and kinematic studies into upper-limb motor control after stroke. Most studies choose movements of the hand or fingers. These tasks are relatively easy to implement under standardized experimental conditions. However, finger tapping performance for instance does not necessarily reflect motor impairments under real world conditions (65), in which object manipulation is an important upper-limb function. Moreover, before being able to manipulate an object, the object needs to be reached. Hence, some authors favor evaluating extension of the elbow, being a main building-block of reaching *(56). Nevertheless, both tasks are equally important, but functionally different, with different levels of complexity and proprioceptive feedback.

In the context of simultaneous kinematic registering, lower-limb fMRI studies present an advantage over upper-limb ones. During brain fMRI, participants are placed deeply within the MRI-tunnel. The more we approach the center of the magnetic field, the more difficult it is to integrate a motion capture system. A system with active markers which emits a signal will be perturbed by the strength of the magnetic field, while passive markers which reflect an emitted light are difficult to see when far in the MRI-tunnel. As the feet often protrude outside of the MRI-tunnel, their motion tracking is easy in contrast to the upper-limb that rests within the MRI-tunnel. For upper-limb tasks, tracking of the end-point (hand/fingers) has been shown the most reliable.

In sum, motion tracking has been performed reliably in both upper and lower-limb protocols, yet with a primary focus on the upper-limb. Both fields have a plentitude of questions that need to be resolved, of which patient stratification on individual brain/behavior characteristics to optimize rehabilitation strategies seems the common denominator. Moreover, the optimal trade-off between ecological valuable movements and their traceability during fMRI still needs to be identified.

### To be or not to be constrained?

In ten studies, participants performed an unconstrained movement in space and time, while the thirteen remaining studies used a predefined pace (auditory signal) or amplitude (straps or orthosis). Post-stroke, motor impairment varies strongly over patients. By using a paced rhythm, task-reproducibility in terms of number of repetitions is indisputably higher. However, the more severe the impairment after stroke, the slower patients seem to move and the more irregular their performance becomes (16). The further a movement is away from the preferred frequency, the higher the costs to perform such a movement (33). Based on the principles of optimal control to move with maximal efficiency at minimal costs (66), an unconstrained movement could thus be more adapted to compare different persons with stroke, with different levels of deficits *(56). We observed that most studies either controlled a fixed movement or monitored the rhythm and amplitude of a “free” movement. When the latter is done with adequate motion tracking systems, it might be preferable because of its higher ecological value, being closer to real live movements with functional relevance - a key factor of successful motor rehabilitation (64).

### How to capture a motor task?

3D motion tracking is the most versatile way to register and analyze human movement, independent of the technology (ultrasonic, optic, electromagnetic) (67). Kinematics extracted from 3D motion tracking systems outside of the MRI that were confronted with activity parameters enabled a fine analysis of brain activity patterns in relation to, for instance, movement irregularity *(22) or compensatory movement intensity *(40). 3D motion tracking during fMRI was documented in 13 of the 23 studies. Of all motion capture methods, optical tracking is recommended for its precise and reliable kinematic analysis after stroke (68). Yet, so far, only one study used optical 3D motion tracking during fMRI post-stroke. This feasibility study, published in 2010 on upper and lower limb movements of one stroke patient *(19), concluded that the kinematic acquisitions were reliable and versatile to enrich fMRI image information, allowing an evaluation of the relationship between functional alterations and brain activations. Still, one case study is not a lot. This underlines again the infancy of the field. Another explanation might be the high cost of a compatible MRI optical motion tracking system. And again, another explanation might be the need for advanced and reliable reconstruction methods. This includes marker-labeling and gap filling to overcome the tracking constraints of optical motion tracking within the MRI-tunnel, like potential skin movement artifacts (69) and data-loss when markers are out of sight. Data-loss is not a problem using rehabilitation-robots like a connected glove or an electro-goniometer, which were used in eight studies. However, these tools capture the movement of the robot rather than the movement of the limb within the robot. Having frequently limited degrees of freedom, directional variations are potentially under-estimated. Using such tools within the MRI is valuable when evaluating the effects of related robot training on brain activity patterns (70) but might be less-representative of the irregular and variable character of movement post-stroke (71). In addition, the sensorial feedback induced by such devices is thought to modify brain activity patterns (38).

Which brings us to the next point that the type of motion tracking tool also impacts the kinematic parameter that can be assessed. The eight studies using a rehabilitation robot, focused mainly on kinematics from the efficiency domain, with only two studies including also a kinematic parameter quantifying movement smoothness. In contrast, although the efficiency domain was equally well represented in 3D motion tracking systems studies, they additionally included variables from different relevant domains, including speed, but also smoothness, planning, accuracy, and posture related kinematics. This is an important advantage of 3D motion tracking as these variables contain valuable information on hemiparetic movement. For instance, movement smoothness is known to be inversely related to the capacity level after stroke (72), and posture related variables contain information about potential compensation strategies (73). Interestingly, recent work by the group of Grefkes *(48) proposed using a “kinematic motor composite score”, based on the principal component explaining the maximal kinematic variance across tasks and participants. The interest of such a composite score is that it may reflect the overall motor performance.

### Coupling fMRI and kinematics

To recall, we identified five different ways kinematic based motion capture has been used in the context of fMRI research after stroke. From simple to more complex, this included: Facilitation of fMRI results interpretation *(41,43,45,52,54,55), optimization of the fMRI contrast paradigm *(19). Guidance of an external device as Transcranial Magnetic Stimulation *(46,47). Provision of motor feedback motor feedback *(21,42,54). And the inference of information about underlying motor control *(18–20,22,39,40,47,49,51). The corresponding results were either obtained using a statistical group analysis, an analysis at the individual level, or by no statistical analysis at all between kinematics and the BOLD signal analysis. In the latter, separated findings were intellectually confronted and discussed. Because of the different quantity and nature of upper and lower-limb studies we will address upper and lower-limb findings separately.

### Upper-limb group level comparisons: understanding motor control

The most significant upper-limb results were found at the group level analysis. In general, kinematic parameters were added as regressors to the second level analysis of the BOLD-signal, which identified regions that varied in activity intensity with the kinematic parameter. For example, Buma et al. *(22), showed that patients with lower levels of hand aperture smoothness during a reach-to-grasp task, recruited additional secondary sensorimotor areas during finger flexion/extension within the fMRI. This was interpreted as a signal of adaptive motor learning strategies to compensate for motor impairments. Interestingly, the jerk being a direct measure of movement quality correlated stronger with brain activation than clinical scales like the Fugl-Meyer Assessment or The Action Research Arm Test. Schaechter & Perdue (2008) *(52), demonstrated that activity in the ipsilateral cortical network was enhanced as a function of task difficulty in stroke patients with good motor recovery. Likewise, Bani-Ahmed et al. *(40) demonstrated the dynamic recruitment of the ipsilateral M1 to be associated with the expression of compensatory trunk use. Hence, ipsilateral M1 was identified as a potential biomarker signaling behavioral compensation. However, methodologically, the tasks within the fMRI (voluntary hand grip force) and outside of the fMRI (reach to grasp) were quite different in nature. This raises the question on the validity of the association between both measures. Ipsilateral M1 activity has previously been related to the control of complex or difficult motor tasks (74). So, could it be that lower grip force and increased trunk compensation are expressions of the same underlying problem: difficulty of motor control after stroke. While the control of hand grip force alone may differ from the control of a reach-to-grasp movement as it includes in addition to muscle force, also correct muscle synergies and intersegment coordination (64,75,76). Evaluation of a comparable task within the fMRI would have been preferable. A direct analysis of movement kinematics was performed by van Dokkum et al. (2018) *(56) who measured the kinematics of an elbow flexion/extension task performed with the less affected upper-limb within the fMRI after stroke. Changes in kinematics were confronted with changes in brain activity patterns, facilitating the latter’s interpretation. Unfortunately, no statistical inference was performed between both measures. Contrarily, Brihmat et al (2020) *(18) did include the normalized amplitude of the passive wrist-extension as covariates in the fMRI analysis. This allowed them to optimize the fMRI paradigm when integrated at the individual level of analysis and to draw a direct link between the activation observed and the task-specific changes in the BOLD signal when integrated at the group level. The latter revealed a direct correlation between the movement amplitude and the primary sensorimotor cortex activity.

### Upper-limb kinematic integration at the individual level: controlling variability

All studies integrating kinematics at the individual level, highlight that it, a) allows to control for differences in task execution within and between subjects *(54), and b) improve activity pattern precision by using kinematics to define the on and offset of the movement block in a blocked fMRI design *(18,19). However, integrating kinematics at the individual level as time-series came at the cost of a decreased signal intensity in the work of Brihmat et al *(18), whereas Casellato et al. (2010) *(19) found an increased and optimized activation map by adding kinematic regressors of an active finger tapping and/ ankle dorsal-plantar flexion task. This difference might be related to the difference in task characteristics: passive regular and paced movement versus active irregular and unconstrained movement. Thus, although seemingly contrasting, both findings were in line with current knowledge. Finger movement with large amplitude elicits significant brain activity, whereas small amplitude movements do not (32). Similarly, the intensity of the BOLD signal is modified by the movement frequency (77). The amplitude and frequency of movement have an important impact on the control of a movement, potentially expressed by other kinematic parameters in the domains of speed, smoothness and accuracy. Notably, when we make fast movements, we have to trade speed for accuracy and this is even stronger for larger movements (78,79), impacting the smoothness of movements (80). By making a distinction between shaping (amplitude and frequency) and structural kinematics, van Dokkum et al. 2017 were able to demonstrate how self-selected pace correlated at the group level with the structure of within movement variability and related to brain activity in the cerebellar-frontal circuit in healthy people. Differently, (81) Shirinbayan et al, (2019) integrated the reaching movement speed at the individual level, as a time-series, after an orthogonalization between the speed regressor and the movement per se, to obtain an independent estimate of the speed related effect. In which movement per se, or its execution was found to be modulated in the primary sensorimotor cortex, in contrast to speed that was rather modulated amongst others in the contralateral premotor cortex and thalamus. The former seems in line with post-stroke studies highlighting the modulation of amplitude to be represented within the primary motor cortex *(18), whereas the latter corresponds with motor control in the frontal-cerebellar circuits following the works of van Dokkum et al. 2017 (82) in healthy participants.

Unfortunately, post-stroke only one study integrated a structural kinematic parameter (i.e other than amplitude or frequency) directly in the fMRI analysis *(51). The angular velocity, resolving from both the amplitude and frequency of movement, was used in the general linear model to investigate rehabilitation induced changes in neuromotor coupling strength. The authors observed a possible relationship between the impairment level and the pattern of brain reorganization. They argued that the combination of kinematics and imaging regresses-out variance in brain activity, accounting for inadvertent motion or inconsistent performance across fMRI sessions. Interestingly, a recent study on 31 Parkinson patients integrated the time course of finger displacement during a finger-thumb opposition task within the fMRI. This model substantially increased cluster size and peak t-values, underlining a marked gain in sensitivity for detecting activation in cortical motor regions. It equally allowed the identification of subcortical areas of this motor loop, e.g., the thalamus and caudate nucleus that would have remained unnoticed otherwise (83). In contrast, by adding kinematics of handwriting at the individual level, and thus correcting for low-level kinematics like writing duration and velocity, a functional specificity in the motor system was observed differentiating between letter and digit writing (84).

### Lower-limb kinematics quantifying rehabilitation gain parallel to fMRI changes

Dobkin and colleagues (2004) *(45) established that the ankle dorsiflexion paradigm was a valuable physiological assay to identify the optimal type, duration and intensity of rehabilitative gait training. Consequently, most subsequent lower-limb studies used the combination of kinematics and imaging to evaluate the effects of various rehabilitation strategies. That is, in chronological order, foot wave tracking produced training effects in both ankle function (kinematics) and brain reorganization (fMRI) *(41) and telerehabilitation with wave tracking showed larger gains in walking capacity than repetitive ankle dorsiflexion movements at self-selected pace *(44). Biofeedback rehabilitation of passive and active ankle dorsiflexion equally modified fMRI parameters and gait, whereby the amount of change in both parameters was strongly correlated *(20). This led the authors to conclude that fMRI is able to capture phases of motor learning after electromyographic biofeedback training. Finally, in a longitudinal pilot study, Gandolla et al (2021) *(46) used kinematics to identify responders to a FES-based therapy, after which between group fMRI modeling was performed to identify the underlying brain organization that may explain why some people do respond to the stimulation and others do not. In all studies, kinematics served only to quantify whether a participant showed functional progress, without being taken into account in the fMRI analysis itself. The only study since Dobkin et al. (2004) *(45) that did not evaluate a rehabilitation technique, evaluated the feasibility of a pedaling motion being a continuous, multi-joint movement rather than the isolated ankle dorsiflexion. The proposed custom-made fMRI compatible pedaling device could indeed be used with fMRI to examine brain activations after stroke *(50). Kinematics, like step length, walking velocity and between legs variables, were used to explain the reduced brain activation volume during pedaling post stroke. The only kinematic parameter that approached significance was the amount of work performed by the paretic limb during pedaling. It may thus not be surprising that the group’s next studies did not explicitly focus on kinematics. Nevertheless, they did show that local and global network connectivity strength was uncorrelated with clinical measures, including the walking velocity (85), but that functional network modifications were larger during active movement than at rest. Yet again, the mean task network connectivity strength was uncorrelated to the corresponding clinical measures (86). Interestingly, they also observed that the Fugl-Meyer Lower Limb score was unrelated to pedaling rate. This makes one wonder whether the right kinematic parameters and the right task were used to unravel the relation between brain network modifications and lower-limb motor function after stroke. Especially when taking into account that pedaling after stroke is characterized by a more variable velocity profile with impaired interlimb coordination and impaired relative limb phasing (87) all these variables were attenuated by the use of a pedaling device with pedal interdependence. This seems to be underlined by the finding that enhanced neural activity during free ankle dorsiflexion (no pedaling device) after stroke correlated with foot-tapping rate outside of the scanner: the stronger the observed neural enhancement, the slower the tapping rate (88). Together it may thus be concluded that integrating pathological relevant kinematics within the fMRI analysis in lower-limb studies after stroke remains by our knowledge largely unexplored. Relevant kinematics representing movement structure should be identified, and evaluated in both fMRI conditions and real-life walking conditions.

### Current challenges and perspectives

Although kinematics has been confronted with brain activity patterns, integrating the kinematic parameters using a multimodal analysis has received comparatively less attention. The multimodal data integration can be defined as a technique that aims to extract information that may not be accessible through a single source, complementing the available information, revealing new information and constraining each other for more reliable output. Motion capture has been primarily used to provide behavior information on how fast and how large movements were executed, either to understand for example how fast movements are performed or to control for differences in movement frequency between participants to understand how the movement per se has been performed. Along-side, when multimodal integration is performed, only simple linear approaches were used, whereas nonlinear relationships between cortical dynamics and movement kinematics can be expected (89, 90).

Interestingly, half of the studies that performed any statistical analysis between kinematics and brain activity, did not assess the movement during fMRI. Although this might have been for practical reasons as MRI compatible mocap is a costly affair, the elephant in the room is whether movements performed outside of the MRI are comparable to those within, when lying down in a physically restraint environment. Is walking velocity functionally related to pedaling speed? How does grip force relate to a reach to grasp task requiring multi-joint coordination? The kinematic quantification of standardized movement assays, as recommended for kinematic upper-limb assessment by stroke rehabilitation experts (14), should be implemented for both upper and lower-limb movements. The subsequent identification of kinematic markers distinguishing neurological recovery versus behavioral compensation can then be confronted with their expression during standardized movements within the fMRI.

Finally, an optimal coupling of fMRI with kinematics requires a short repetition time and an efficient 3D motion MRI-compatible tracking device. In an ideal world the sample frequencies of both modalities should be comparable. Yet, although imaging sample frequency is improving through imaging techniques like multiband fMRI (91), current repetition times vary between one and three seconds. This is far from the optimally recommended sample frequency for motion capture, to know 100 Hz (14). Although the latter may be discussed, as all measured body movements are contained within frequency components under 20 Hz (92), thus valuable information may equally be obtained with lower sample frequencies. Correlating the evolution of both time-series requires the down-sampling of the kinematic time series which may induce information loss, especially in the case of repetitive motion. For example, when the timing of the imaging volume coincides systematically with the zero-velocity turning point of the rhythmical motion, a false representation of movement is created. Approaches known in human motor control studies like the Nyquist-Shannon sampling theorem might thereby provide alternative solutions limiting information loss (93). Another related challenge that merits further exploration is the link between the fast fluctuations of movement time series vs the slow and stable BOLD response that is maintained over rhythmic motion in a blocked fMRI design.

Thus, even though the value of combination kinematics with brain imaging has been underlined repetitively, and seems mandatory to improve our understanding of brain plasticity over recovery in a holistic manner with increasing reliability, certain recommendations seem in order. Notably:

- Performing direct motion capture during fMRI, using minimally restricted motion capture devices like optical motion tracking.
- When direct motion capture is not possible, task correspondence within and outside the MRI should be maximized.
- Analyzing both shaping and structural kinematics, covering all kinematics domains.
- Using high field MRI with the lowest repetition time possible.
- At the individual level fMRI analysis, kinematics should be more explored as a time-series.
- Exploring non-linear relationships between kinematics and brain activity patterns.

Of course, these recommendations are based on the English literature of MRI motor function studies after stroke. We recognize that there is broad range of other techniques available to measure brain activity, such as Electroencephalography (EEG), Magnetoencephalography (MEG), Near Infrared Spectroscopy (NIRS) or Positron Emission Tomography (PET). Yet, a recent review on motion capture and EEG came to comparable conclusions (94). Also, these techniques have limited spatial resolution and do not allow the exploration of subcortical structures, whose role during finger tapping was unveiled by the multimodal integration of kinematics and brain imaging (83). We also recognize the use of the joint approach in both healthy volunteers and other pathologies. Literature from other fields has been searched and integrated into the discussion.

## Conclusion

The present review has explored studies related to the combination of kinematics with fMRI in the study of post-stroke motor function. The reported outcomes suggest that simultaneous use of these two techniques permits to obtain novel information compared to classical fMRI analysis. Quantifying movement kinematics and including them in the fMRI analysis at individual or group level has been proven crucial for interpreting the fMRI results, in order to link the observed differences to differences in movement parameters, occurrence of involuntary movements, effect of rehabilitation intervention, recovery, or underlying motor control. Although the field is moving forward, as highlighted by recent studies in healthy participants and domains out of stroke, the current state of the art emphasizes the need to both control and report movement scaling kinematic like frequency and amplitude and to include structural kinematics into the multimodal analysis. Kinematics used as a covariate of no-interest allows to control for within and between subject variability, allowing to draw conclusions on the execution of a motor task per se. Whereas as covariate of interest it may highlight the neural substrates of this variability, explaining potentially deficit related patterns in the context of neurorehabilitation.

Nevertheless, the multimodal coupling of kinematics and imaging is still subject to practical and technological limitations. Further research is required to optimize and standardize both data measurement and treatment procedures, improving the overall quality of related studies. We believe that our findings and recommendations might enable the field to move forward and fully use the potential of a multimodal kinematic imaging approach to unravel the complexity of post-stroke motor function and recovery.

## Declarations

### Ethics approval and consent to participate

Not applicable.

### Consent for publication

Not applicable.

### Availability of data and materials

Additional data that would not appear in the review are available upon reasonable request.

### Author contributions

ZB carried out the paper selection and data extraction process with validation of LvD and ELB. ZB and LvD rated methodological quality and wrote the article. All authors revised the article for publication.

## Data Availability

All data produced in the present study are available upon reasonable request to the authors.

## Appendix

### Appendix A: Selection by search terms

| **Term of interest** | **Number of results** | **Motor engine** | **Date of search** |
| --- | --- | --- | --- |
| fMRI | 260542 | 4 databases | 17/08/2022 |
| + Stroke | 17700 | 4 databases | 17/08/2022 |
| + motor control | 3766 | 4 databases | 17/08/2022 |
| + motion tracking | 199 | 4 databases | 17/08/2022 |

## Abbreviations

fMRI: functional Magnetic Resonance Imaging
Mocap: Motion capture
EEG: Electroencephalography
MEG: Magnetoencephalography
fPET: functional Positron Emission Tomography
fNIRS: functional Near-Infrared Spectroscopy
TMS: Transcranial Magnetic Stimulation
VR: Virtual Reality

